# Modeling the impact of social distancing, testing, contact tracing and household quarantine on second-wave scenarios of the COVID-19 epidemic

**DOI:** 10.1101/2020.05.06.20092841

**Authors:** Alberto Aleta, David Martín-Corral, Ana Pastore y Piontti, Marco Ajelli, Maria Litvinova, Matteo Chinazzi, Natalie E. Dean, M. Elizabeth Halloran, Ira M. Longini, Stefano Merler, Alex Pentland, Alessandro Vespignani, Esteban Moro, Yamir Moreno

## Abstract

The new coronavirus disease 2019 (COVID-19) has required the implementation of severe mobility restrictions and social distancing measures worldwide. While these measures have been proven effective in abating the epidemic in several countries, it is important to estimate the effectiveness of testing and tracing strategies to avoid a potential second wave of the COVID-19 epidemic. We integrate highly detailed (anonymized, privacy-enhanced) mobility data from mobile devices, with census and demographic data to build a detailed agent-based model to describe the transmission dynamics of SARS-CoV-2 in the Boston metropolitan area. We find that enforcing strict social distancing followed by a policy based on a robust level of testing, contact-tracing and household quarantine, could keep the disease at a level that does not exceed the capacity of the health care system. Assuming the identification of 50% of the symptomatic infections, and the tracing of 40% of their contacts and households, which corresponds to about 9% of individuals quarantined, the ensuing reduction in transmission allows the reopening of economic activities while attaining a manageable impact on the health care system. Our results show that a response system based on enhanced testing and contact tracing can play a major role in relaxing social distancing interventions in the absence of herd immunity against SARS-CoV-2.

---

The first report of a new infectious disease, later coined COVID-19, appeared on 31 December 2019^1^. As of 2 May 2020, the virus has spread to 187 countries with more than 3.4 millions confirmed cases worldwide, and killing more than 240,000 people^2^. As the number of confirmed COVID-19 cases increased and the expansion of the disease entered into a global exponential growth phase, a large number of affected countries were forced to adopt non-pharmaceutical interventions at an unprecedented scale. Given the absence of specific antiviral prophylaxis, therapeutics or a vaccine, non-pharmaceutical interventions ranging from case isolation and quarantine of contacts, to the lock-down of entire populations have been implemented with the aim of suppressing/mitigating the epidemic before it could overwhelm the health care system. Although these aggressive measures appear to be successful in reducing the number of deaths and hospitalizations^3,4^, and in reducing the transmission of the SARS-CoV-2 virus, the absence of herd immunity after the first wave of the epidemic points to a large risk of resurgence when interventions are relaxed and societies go back to a “business as usual” lifestyle^5-7^. It is therefore of paramount importance to analyze different mitigation and containment strategies aimed at minimizing the risk of potential additional waves of the COVID-19 epidemic while providing an acceptable trade-off between economic and public health objectives.

In the present work, through the integration of anonymized and privacy-enhanced data from mobile devices and census data, we build a detailed sample of the synthetic population of the Boston metropolitan area in the United States (see Figure 1a and 1b). This synthetic population (Figure 1a) is used to define a data-driven agent-based model of SARS-CoV-2 transmission and to provide a quantitative analysis of the evolution of the epidemic and the effectiveness of social distancing interventions. The model allows us to explore strategies concerning the lifting of social distancing interventions in conjunction with testing and isolation of cases and tracing and quarantine of exposed contacts. Our results indicate that after the abatement of the epidemic through the “stay at home” orders and halt to all nonessential activities, a proactive policy of testing, contact tracing and contacts’ household quarantine allows the gradual reopening of economic activities and workplaces, with a low COVID-19 incidence in the population and a manageable impact on the health care system.

**Figure 1.**
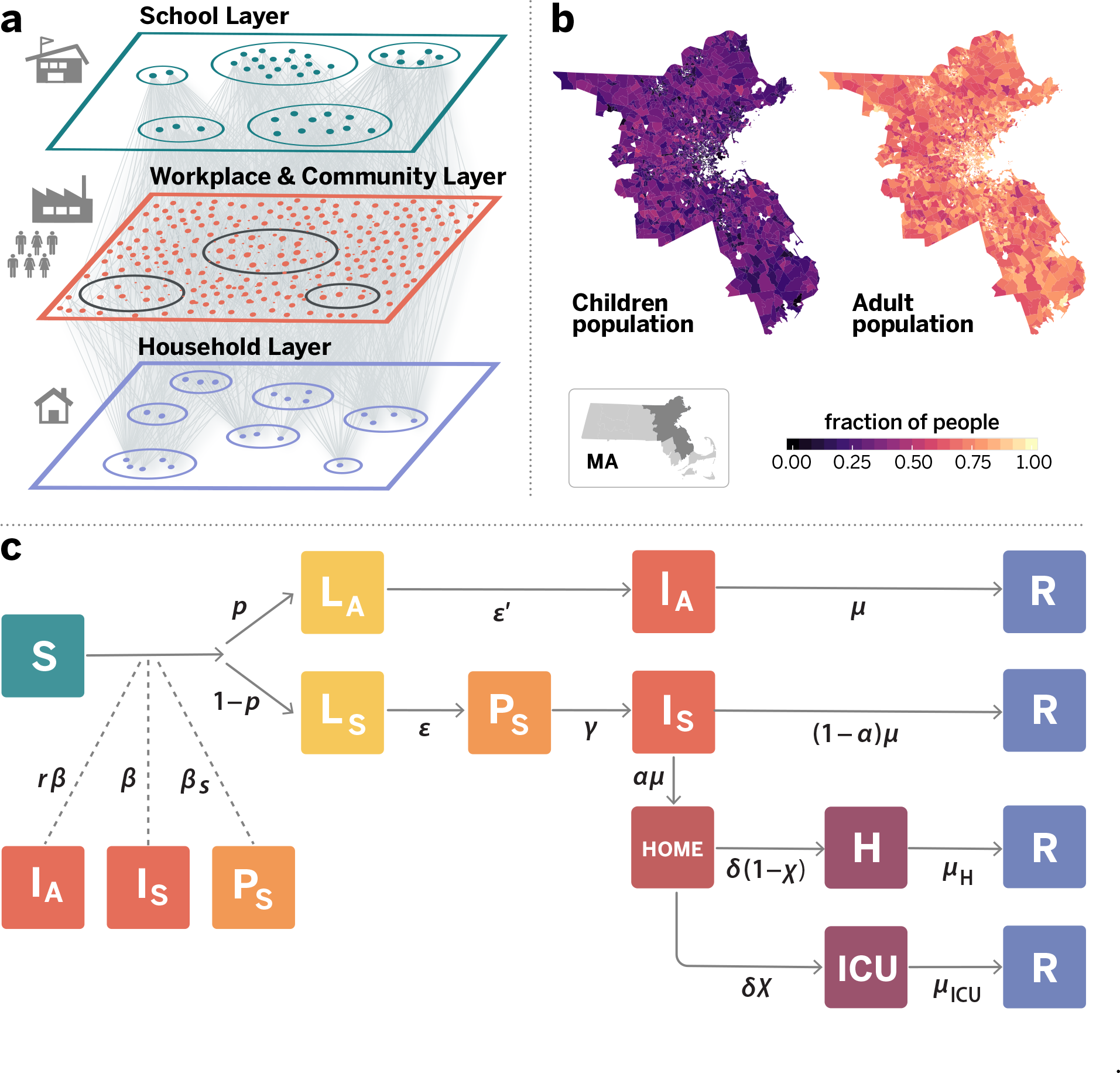
Model components. Panel **a** is a schematic illustration of the weighted multilayer synthetic population built from mobility data in the metropolitan area of Boston. The agent-based system is made up by around 64000 adults and 21000 children, whose geographical distributions are shown in panel **b**. Nodes are connected by more than 5 million weighted edges. Community layers (that include workplaces), are further classified into categories according to Foursquare’s taxonomy of places. Panel c displays the compartmental model used to describe the natural history of the disease as well as the transition rates between the different states. Specifically, we consider Susceptible (S), Latent asymptomatic (L_A_), Latent symptomatic (L_S_), Pre-symptomatic (P_S_), Infectious asymptomatic (I_A_), Infectious symptomatic (I_S_), Hospitalised (H), Hospitalized in intensive care (ICU) and Recovered (R) individuals. More details of the model and the transitions between compartments are provided in Methods and the SM.

To provide a quantitative estimate of the contact patterns for the population of agents and to build the synthetic population of the Boston Metropolitan Area (BMA), we used detailed mobility and socio-demographic data and generated a network that encodes the contact patterns of about 85,000 agents in the area during a period of six months (see Supplementary Material, SM). Agents are chosen to be representative of the different census areas in the Boston area following the methodology used in Ref. ^8^. This defines a weighted multilayer network consisting of three layers representing the network of social interactions at (1) workplace/community level (W+C), (2) households, and (3) schools, as shown in Figure 1a. Connections between two agents in the W+C layer are estimated from the data by the probability of both being present in a specific place (e.g. restaurant, workplace, shopping) weighted according to the time they have spent in the same place. A second layer represents the households of each anonymous individual. Using the home census block group of each anonymous user we associate each individual to a specific household profile based on socio-demographic data at US census block group level^9^. Families are generated by randomly mixing nodes from the community living in the same census block group, following the statistical features of family types and sizes. Finally, a third layer represents the contacts in the schools (i.e., every node represents one synthetic student and has contacts only with other individuals attending the same school). To study the evolving dynamics of the infection, we implemented a stochastic, discrete-time compartmental model in which individuals transition from one state to the other according to key time-to-event intervals (e.g., incubation period, serial interval, and time from symptom onset to hospital admission) as from available data on SARS-CoV-2 transmission. The natural history of the disease is captured by the epidemiological model represented in Figure 1c, where we also show the transition rates among compartments^8,10-12^. The model considers that susceptible individuals (S) become infected through contact with any of the infectious categories (infectious symptomatic (I_S_), infectious asymptomatic (I_A_) and pre-symptomatic (P_S_)), transitioning to latent compartments (L_S_) and (L_A_), where they are infected but not infectious yet. Latent individuals branch out in two paths according to whether the infection will be symptomatic or not. We also consider that symptomatic individuals experience a pre-symptomatic phase and that once they develop symptoms, they can experience diverse degrees of illness severity, from mild symptoms to being hospitalized (H) or in need of an intensive care unit (ICU)^13^. Finally, individuals transition in the removed compartment (identifying recovered or dead individuals). The model assumes a basic reproductive number *R*_0_ = 2.5, which together with the rest of the parameters (see SM, table S1) yields a generation time *T_g_* = 6.6 days. We consider a 25% fraction of asymptomatic individuals. We report the full set of parameters used in the model and an extensive sensitivity analysis in the SM file. The model is not calibrated to account for the specific evolution of the COVID-19 epidemic in Boston as it is aimed at showing the effect of different non-pharmaceutical interventions rather than providing a forensic analysis of the outbreak in the BMA. Details on the generation of the synthetic population network and the infection transmission model are provided in the SM.

## Results

To provide a baseline of the COVID-19 impact in the Boston metropolitan area, we have first investigated an unmitigated scenario in which no interventions are implemented. Results for the unmitigated scenario are shown in Figure 2, panels a-c. A COVID-19 unmitigated epidemic would have a peak incidence of 25.2 (95% C.I: 23.8-26.4) newly infected individuals per 1,000 people. The epidemic follows a typical trajectory, namely, when the effective reproduction number *R_t_* as a function of time (panel c) becomes smaller than 1, the transmission dynamics slows down and eventually vanishes after having infected about 75% of the population (Figure 2b). Figure 3a shows the evolution of the number of new severely affected patients who require hospitalization and admission into ICUs. At the peak of the unmitigated epidemic, the number of ICU beds needed exceeds by far the available capacity (dashed horizontal line in Figure 3a) by more than a factor of 12, thus indicating that the health care system would suffer large service disruptions, resulting in additional deaths due to hospitals overcrowded with patients with COVID-19^14^. It is worth noting that current fatality rates consider the general availability of ICU beds and critical care capacity, if this would not be possible the fatality rate may increase dramatically.

**Figure 2.**
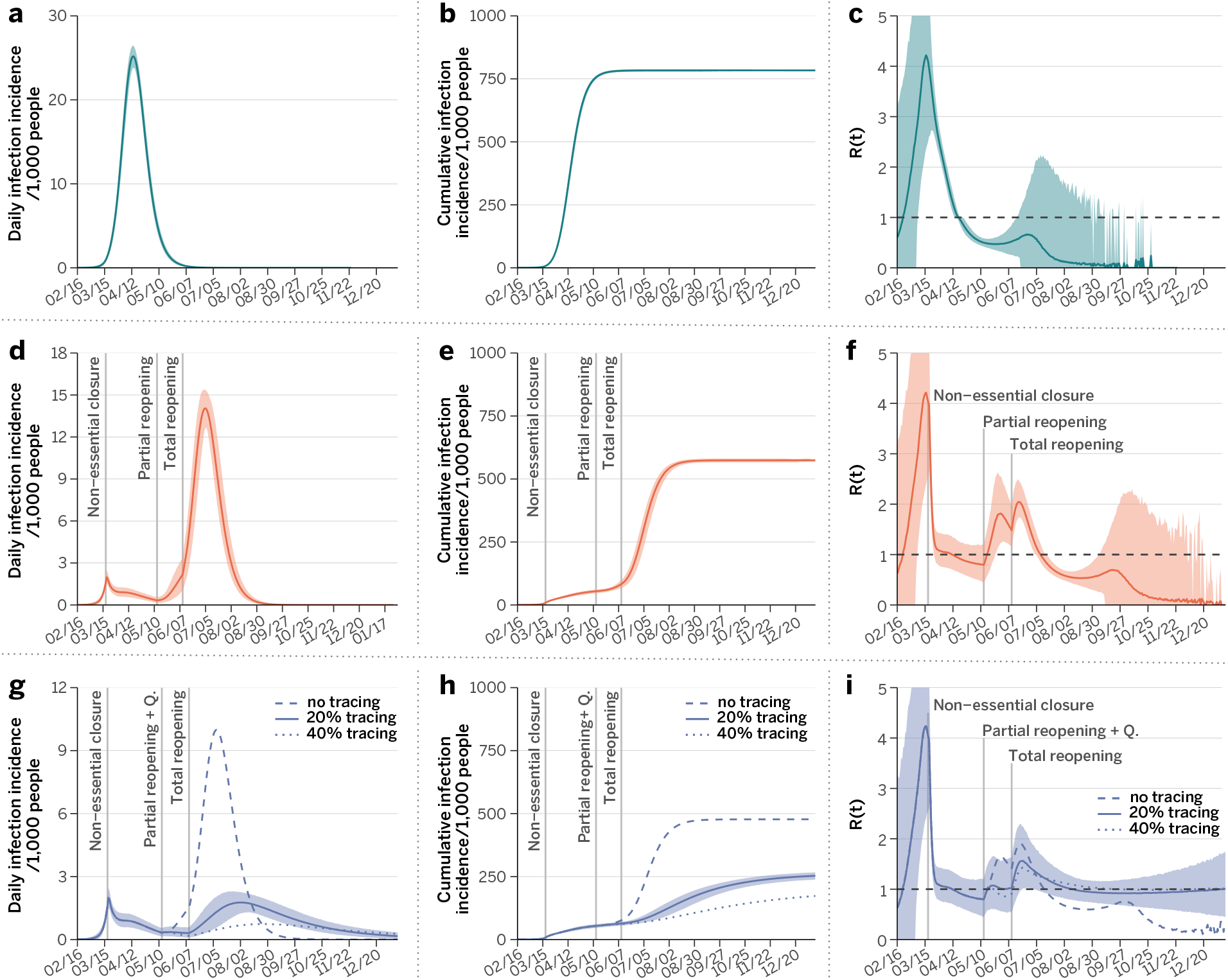
Impact of COVID-19 under different scenarios. Evolution of the number of new cases (**a, d, g**), the outbreak size (**b, e, h**) and the effective reproductive number (**c, f, i**) as a function of time in each situation studied. Results of the SARS-CoV-2 transmission dynamics are shown for the unmitigated scenario (top panels **a-c**), and the two social distancing interventions considered, LIFT (**d-f**) and LET scenarios (**g-h**). In both cases, we considered the closure of schools and non-essential places for 8 weeks. This is the strictest lock-down period, which is followed by a partial lifting of the stay-at-home policy whose duration is set to 4 weeks. During the partial lifting, all places in the community layer are open except mass-gathering locations (restaurants, theaters, etc, see SM). Finally, a full reopening takes place after the period of partial lifting ends (relevant events are marked with the vertical lines). Panels **d-f** consider that no other measures are adopted concurrently to the lifting of the restrictions, whereas the results in panels **g-i** have been obtained when the reopening is accompanied by an active policy consisting of testing the symptomatic individuals, home isolating them, and quarantining their household and the households of a fraction of their contacts, as indicated in the legend of the bottom panels. Note that the vertical scales of panels **a, d**, and **g** are not the same and that both the number of new cases and total cases are per 1,000 inhabitants. In all panels the solid line represents the average over 10,000 simulations and the shaded region the 95% C.I.

**Figure 3.**
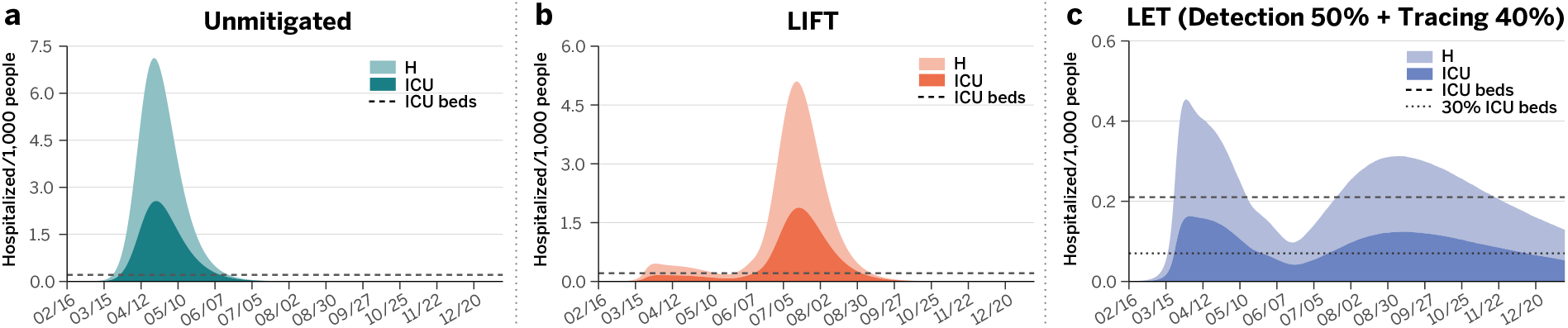
Impact on the Boston health care system. Estimated number of individuals per 1,000 inhabitants that would need hospitalization (H), and intensive care (ICU) for each of the three scenarios considered in Figure 2. Panel a corresponds to the unmitigated situation, whereas results for the LIFT and LET strategies are shown in panels b and c, respectively. The horizontal dotted-dashed lines represent the ICU basal capacity of the Boston health care system. The dotted line in panel c indicates 30% of the ICU basal capacity.

To avoid the harmful effects of an unmitigated COVID-19 epidemic, governments and policy makers across the world have relied on the introduction of aggressive social distancing measures. In the United States, as of April 15 2020, it was estimated that more than 95% of the population was under a “stay at home” or “shelter in place” order^15,16^. To model the social distancing policies implemented in the whole Boston metropolitan area, we have considered March 17, 2020 as the average starting date of social distancing policies that include school closures, the shut down of all non-essential work activities as well as mobility restrictions (see SM for details). This scenario mimics the social distancing intervention implemented in most of the high income countries in Europe and across states in the US. Such extreme social distancing policies come with very large economic costs and social disruption effects^17^, thus prompting the question of what exit strategy can be devised to restart economic activities and normal societal functions^18^. For this reason, we explore two different scenarios for lifting social distancing interventions:

• Lift scenario (LIFT): the “stay at home” order is lifted after 8 weeks by re-opening all work and community places, except for mass-gathering locations such as restaurants, theaters, and similar locations (see SM). We also assume that symptomatic COVID-19 cases are isolated within 2.5 days. The latter partial re-opening is enforced for another 4 weeks, which is followed by a full lifting of all the restrictions that remained. We consider that schools will remain closed given the impending summer break in July and August, 2020. In fact, some school systems, like the Boston Public schools, have announced they will remain closed through the 2019-2020 school year.
• Lift and enhanced tracing (LET) scenario: The “stay at home” order is lifted as in the previous scenario. Once partial reopening is implemented, we assume that 50% of symptomatic COVID-19 cases can be tested for SARS-CoV-2 infection, on average, within 2 days after the onset of symptoms and that they are isolated at home and their household members are quarantined successfully for 2 weeks (a sensitivity analysis for lower rate of isolation and quarantine is presented in the SM). We also assume that a fraction of the non-household contacts (we show results for 20% and 40%) of the symptomatic infections can be traced and quarantined along with their household as well — note that we consider that the contact tracing is more likely to pick up interactions proportionally to the time spent together.

The above scenarios are mechanistically simulated on the multilayer network of Figure 1a, by allowing different interactions (between effective contacts) according to the simulated strategy. As a result, the average number of interactions in the W+C layer goes from 10.86 (95% C.I.:1.51- 42.39) under the unmitigated scenario, to 4.10 (95% C.I.:0-23.79) for the partial lock-down and only 0.89 (95% C.I.: 0-8.39) contacts for the stay at home policy, see Methods and SM for more details. This result is in agreement with previously published work^19^ and recent reports in the New York City area^20^. It is worth remarking that the fluctuations in the number of contacts in the stay at home order is due to a large extent to contacts that take place in grocery stores and other essential venues.

The numerical results show that the LIFT scenario, while able to temporally abate the epidemic incidence, does not prevent the resurgence of the epidemic and a second COVID-19 wave when the social distancing measures are relaxed. In Figure 2d, we show that following the lifting of social distancing the infection incidence starts to increase again, and the effective reproductive number, that dropped by circa 75% and reached values below 1 with the intervention, increases to values up to 2.05 (95%CI: 1.73-2.47) (see Figure 2f). Indeed, at the time of lifting the social distancing intervention the population has not achieved the level of herd immunity that would protect it from the resurgence of the epidemic. We also estimate that a second wave of the epidemic still has the potential to infect a large fraction of the population (Figure 2e) and to overwhelm the health care systems, as shown in Figure 3b. The number of ICU beds needed, although half the unmitigated scenario, is still exceeding by far the estimated availability^5-7,21^. Such scenario would imply resorting again to major distancing policies, as it would be untenable to let run the epidemic again. This suggests that lifting social distancing without the support of additional containment strategies is not a viable option.

In the case of the LET scenario, the lifting of the social distancing intervention goes along with a significant amount of contact tracing and precautionary quarantine of potentially exposed individuals. The quarantine is not limited to the contacts of the identified symptomatic COVID-19 case, but extended to their households. This strategy amounts to a simplified tracing of contacts of contacts, that would not require extensive investigations within households. In other words, this strategy does not require the tracking of single individuals but considers households as the basic unit. Households could be monitored though, with daily calls or messages to ascertain the onset of symptomatic infections, and provide medical support as needed.

Figure 2g shows results obtained for different levels of tracing (no tracing, 20% and 40%) of the contacts of the symptomatic isolated COVID-19 cases. By comparing Figure 2d with Figure 2g (for no tracing), we find that quarantining households of symptomatic subjects alone is not enough to significantly change the course of the epidemic and the conclusions reached for the first of these scenarios. When 40% or more of the contacts of the detected symptomatic infections are traced and they and their households quarantined, the ensuing reduction in transmission leads to a noticeable flattening of the epidemic curve and appears to effectively limit the possible resurgence of a second epidemic wave. It is also worth noticing that we assume the absence of other additional and minimally disruptive social distancing policies such as crowd control, smart working, wearing of masks, etc., that could lead to a further reduction of the transmissibility of the virus with respect to our estimations. It is important to stress that the contact tracing proposed here works at the level of household unit, simplifying also the monitoring and follow up process, by contacting only one member of the household to monitor the onset of symptoms among all members (we further explore other isolation/quarantine strategies in the SM). Figure 3c and Table1 show the burden in hospitalization and ICU demand in the unmitigated situation and the two mitigation scenarios. The LET scenario allows relaxation of the social distancing interventions while maintaining the hospital and ICU demand at levels close to the health-care availability and surge capacity. For the sake of completeness in the SM file we report the analysis for a LIFT scenario including the school and University reopening in the fall. The results show that in absence of additional containment policies the tracing effort would need to be raised of about 50% to cope with the increased number of infections.

## Discussion

The efforts in the suppression and mitigation of COVID-19 are pursuing the objectives of preserving the health care system from disruptive failures due to overwhelming stress imposed by the large number of severe cases, and of minimizing the morbidity and mortality related to the epidemic. The aggressive social distancing interventions implemented by many countries in response to the COVID-19 pandemic appear to have achieved the interruption of transmission and the abatement of the epidemic, although at the price of huge societal disruption and economic costs. In such a context, the identification of “exit strategies” that allow restarting economic and social activities while still protecting the healthcare systems and minimizing the burden of the epidemic is of primary importance. Several modeling studies have already pointed out that resuming economic activities and social life is likely to lead to a resurgence of the COVID-19 epidemic, and combined social distancing interventions of different degrees and intensity have been proposed to substantially delay and mitigate the epidemic^17,21^. These interventions should be maintained for long periods of time and still generate economic loss and widespread disruption to social life. Here we show how testing, contact tracing strategies at scale, based on home isolation of symptomatic COVID-19 cases and the quarantine of a fraction of their contacts’ household, has the potential to provide a viable course of action to manage and mitigate the epidemic when social distancing interventions are progressively lifted^22 23^. These strategies present us with logistic challenges that include large-scale and rapid diagnostic capacity, and a large surge in the number of contact tracers. We have investigated what fraction of the population would be isolated/quarantined under the proposed contact tracing and isolation strategy. Figure 4a shows the fraction of households that needs to be quarantined. Assuming the identification of 50% of the symptomatic infections, and tracing of 40% of their contacts and households, only about 9% of the population would be quarantined at any time. While this is certainly a relevant fraction of the population, it is a much better option if compared with massive social distancing policies affecting the entire population that last for months.

**Figure 4.**
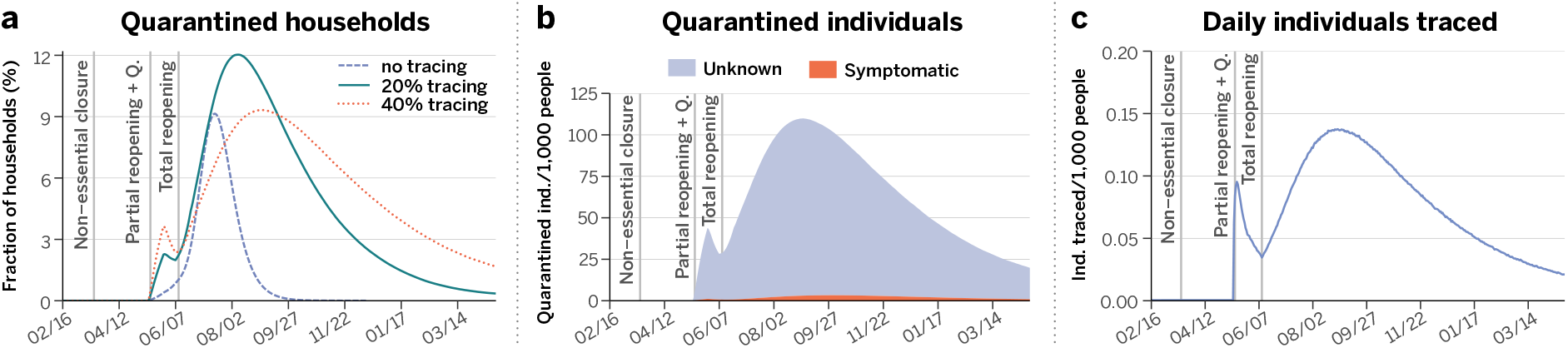
Affordability of the best way-out scenario. LET strategy with 50% detection and 40% tracing. (**a**) Fraction of the population that needs to be put under quarantine as a function of time and percentage of contact tracing. (**b**) Health status of the individuals that are quarantined for a contact tracing level of 40%. Note that only symptomatic individuals are tested, which implies that a large fraction of the quarantined population is of unknown status. This fraction of individuals quarantined with unknown health condition could be reduced if the capacity to do more tests increases. As it is shown, the pandemic might span over several months depending on the level of contact tracing. (**c**) Number of individuals whose contacts are traced each day per 1,000 persons. Relevant intervention actions are signaled by the vertical dashed lines in all panels.

In Table 1, we report the number of symptomatic infections for which the contact tracing investigation should be performed in the basic scenarios. This number provides an estimate of the contact tracers per 1,000 individuals. It is important to note that the more effective is the contact tracing starting from each individual, the smaller is the number of generally traced households because the epidemic has lower incidence rates. In addition, as illustrated in Figure 4b, the health status of the vast majority of quarantined individuals is unknown as contact tracing does not imply testing. The curves in Figure 4a constitute the upper bounds for each simulated case. If we assume that the capacity to do massive testing would likely ramp up in the near future, then it is expected that the actual number of people in quarantine could be significantly lowered by testing the household. This would also alleviate the burden on household members that could not go to work and increase compliance of isolation for the positive cases. It is also worth remarking that many of the logistic challenges faced with massive contact tracing might possibly be eased by digital technologies that are currently being investigated across the world following the examples of COVID-19 response in Asian countries^23^. Also it may be difficult to quarantine the entire household of individuals who are potentially exposed, since this is a hardship suffered with great uncertainty about their risk of infection. Offering other logistic quarantine solutions (quarantine centers, hotel rooms) might significantly raise the rate of compliance.

**Table 1:**
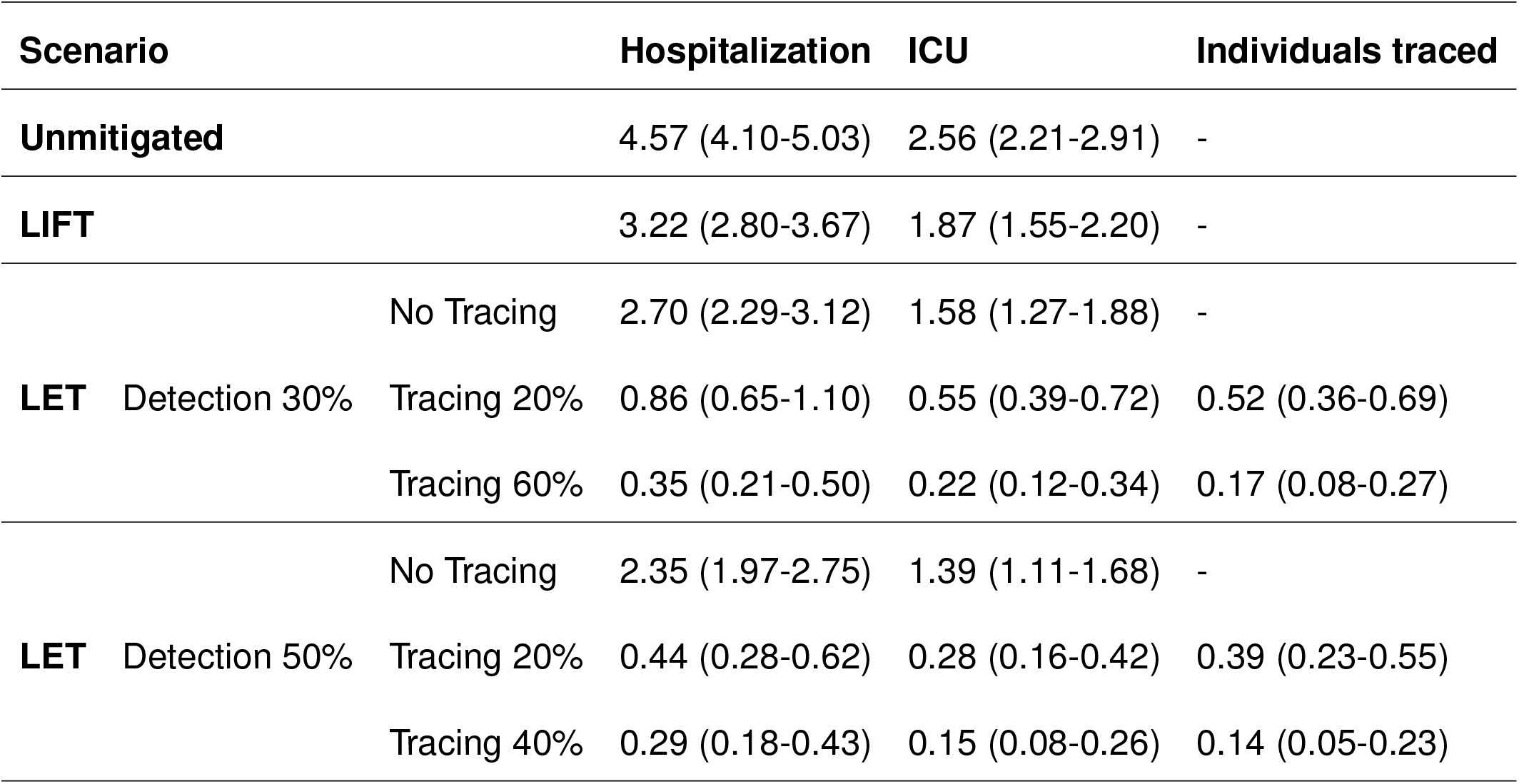
Mean and 95% C.I. of the number of normal hospitalizations, ICU hospitalizations and symptomatic individuals identified/traced (when applicable) at the peak of the epidemic per 1,000 people. The estimated availability of ICU beds is 0.21 beds per 1,000 people.

These results were obtained under several assumptions. There are very large uncertainties around the transmission of SARS-CoV-2, in particular, the fraction of sub-clinical and asymptomatic cases and their transmission. Estimates of age-specific severity are informed from the analysis on individual-level data from China and other countries, and subject to change as more US data become available. We also do not include specific co-morbidities or pre-existing conditions of the specific BMA population. For this reason, in the SM we perform an extensive sensitivity analysis showing that the modeling results discussed here are robust to the plausible range of parameter values for the key time-to-event intervals of COVID-19 (e.g., incubation period, serial interval, and time from symptom onset to hospital admission, etc.). We are also not considering potential changes to the virus transmissibility due to environmental factors, in particular, seasonal drivers such as temperature and humidity. The modeling does not consider possible reintroduction of SARS-CoV-2 in the population from infected travelers. Strategies based on testing, isolation and contact tracing will eventually fail in the presence of a large number of case importations, thus travel restrictions and screening may need to be introduced to/from places that show sustained local transmission. Finally, we did not include the effect of the widespread use in the population of masks or other personal protective equipment. These active protection measures could contribute to the reduction of transmissibility and improve the effectiveness of the exit strategies modeled here.

The modeling of the impact of testing, contact tracing and isolation on second-wave scenarios of the COVID-19 epidemic could be instrumental to national and international agencies for public health response planning. Our results indicate that gradually removing the restrictions imposed by social distancing could lead to a second wave with the potential to overwhelm the health care system if not combined with strategies aimed at the prompt testing of symptomatic infections and the tracing and quarantine of as many of their contacts as possible. While we show that contact tracing and household quarantine at scale may be effective even assuming a complete lifting of the social distancing measures, future decisions on when and for how long to relax policies will need to be informed by ongoing surveillance. For instance, smart working from home for people who can adhere to it without serious disruptions should be encouraged. This, as well as other minimally disturbing policies together with transition periods in which the partial lift of social distancing goes along with efficient and large-scale testing, contact tracing and monitoring of the epidemic should be considered in the definition of exit strategies from large scale “stay at home” orders. Important enough, as shown in the SM, isolation of symptomatic infectious individuals within their households is a valid strategy only when the whole household is also set in quarantine. As such, providing spaces where symptomatic people can be individually isolated would help to reduce the burden of these measures on the population.

## Methods

### Weighted synthetic population

Our synthetic population is constituted by circa 85000 nodes, of which 64000 are adults and 21000 correspond to children (defined as individuals aged 17 or less), see Figure 1b. The total number of interactions among these individuals before social distancing is given by more than 5M edges, see SM for a finer description.

#### Community weighted contact network

The community network is approximated using 6 months of data observation in the Boston area from anonymized users who have opted-in to provide access to their location data anonymously, through a GDPR-compliant framework provided by Cuebiq. Individuals performing the analysis were legally required to never single out identifiable individuals and not make attempts to link these data to third party data about an individual. In this layer each agent in our synthetic population represents an anonymous individual of the real population. The data allow us to understand how infection can propagate in each layer by estimating co-location of two individuals in the same setting. We use a large database of 83000 places from Foursquare API in the BMA. Specifically, the weight, 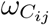, of a link between individuals *i* and *j* within the workplace plus community layer is computed according to the expression:

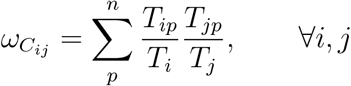

where *T_ip_* is the total time that individual *i* was observed at place *p* and *T_i_* is the total time that individual *i* has been observed at any place set within the workplace plus community layer. Note that while the mobility data set we use is large, co-location events between individuals are still quite sparse. Because of this sparsity, and to protect individual privacy in our analysis, we have adopted this probabilistic approach to measure co-presence in all locations mapped in the dataset. Since agents are representative of the different census areas and groups of the Boston area, our probabilistic approach is a good proxy for the real probability of co-presence between those groups/areas when networks are scaled up to the total population of the Boston area, that is approximately 4,628,910 inhabitants. Finally, for robustness and computational reasons, we have included only links for which 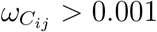.

#### Household weighted contact network

We first localize individuals’ approximate home place according to the US census block group. Then we assign a type of household based on Table B11016: Household Type by Household Size from US Census 2018^9^, and mix individuals that live in the same block according to statistics of household type and size. Finally, children are assigned to households as described in the main text. We also assign individuals an age group based on Table B01001: Sex and age from the US Census 2018. To assign weights, we assume that the probability of interaction at a household is proportional to the number of people living at the same household (well-mixing). Therefore, the weight, 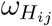, of a link between individuals *i* and *j* within the same household is given by:

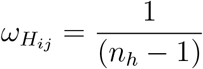

where *n_h_* is the number of household members. This fraction is assumed to be the same for all individuals in the population.

#### School weighted contact network

To calculate the weights of the links at the school layer, we mix together all children that live in the same school catchment area. Interactions are considered well-mixed, hence, the probability of interaction at a school is proportional to the number of children at the same school. Therefore, the weight, 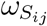, of a link between children *i* and *j* within the same school is given by:

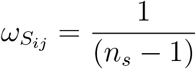

where *n_s_* is the number of school members.

### Calibration of intra-layer links

Initially, we need to calibrate layer weights. We rescale the weight of all links in each layer so that the average number of daily contacts matches the estimation provided in Ref.^24^ based on the analysis of contact survey data from 9 countries^25-28^. In particular, we estimate that in the unmitigated scenario the number of daily contacts is 10.86, 4.11 and 11.41 in the community+workplace, household and school layers, respectively (see SM).

### Stochastic simulations of the COVID-19 dynamics

We describe the COVID-19 transmission process using a discrete time Susceptible-Latent-Infected-Removed (SLIR) stochastic model, with some extra compartments to incorporate the special characteristics of SARS-CoV-2 infection, Figure 1c. In particular, at each time-step *t* (1 day), the infectious asymptomatic (I_A_), infectious symptomatic (I_S_) and pre-symptomatic (P_S_) individuals can transmit the disease to susceptible (S) subjects with probability *rβ*, *β* and *β_S_*, respectively. If the transmission is successful, the susceptible node will move to the latent asymptomatic state (L_A_) with probability p or to the latent symptomatic state (L_S_) with probability (1 – *p*). A latent asymptomatic individual becomes infectious asymptomatic after a period (*ϵ′*)^-1^, whereas latent symptomatic subjects transition, after a period *ϵ*^-1^, to the pre-symptomatic (P_S_) compartment. The average period to develop the disease and move to the infectious symptomatic state is *γ*^-1^. Infectious asymptomatic nodes will be removed (R) after an average of *μ* steps. Conversely, infectious symptomatic nodes can either recover after that period with probability (1 – *α*) or, with probability *α*, these nodes will need hospitalization. It is considered that due to their symptoms they will self-isolate at home after an average period of *μ*^-1^. Then, depending on the severity of the symptoms, after a period *δ*^-1^ the hospitalization will be normal with probability (1 – *χ*) or require ICU care with probability *χ*. Finally, individuals that are either hospitalized or at ICU become removed with probability *μ*_H_ or *μ*_ICU_, respectively. We initialize the model in the city of Boston by selecting an attack rate on the 17th of March of 1.5% (a sensitivity analysis of this quantity is provided in the SM).

### Social distancing strategies.

To simulate social distancing measures, we modify the synthetic population such that:

- School closures are simulated by removing all the schools from the system simultaneously.
- Partial “stay at home”. It assumes that all places are open except restaurants, nightlife and cultural places. Closures of these places are simulated by removing the interactions that occur in any place that falls into that category according to Foursquare’s taxonomy of places. This is the situation after the first reopening.
- Full lock-down and confinement, namely, schools and all non-essential workplaces are closed. Here we close all workplaces except essential ones and remove interactions that occur at them. Essential workplaces are: Hospitals, Salons, Barbershops, Grocery Stores, Dispensaries, Supermarkets, Pet Stores, Pharmacies, Urgent Care Centers, Dry Cleaners, Drugstores, Maternity Clinics, Medical Supplies and Gas Stations.

The connectivity distributions for each of the scenarios simulated as well as other statistics related to the effects of the lock-down are shown in the SM.

## Data Availability

Under request.

## Acknowledgements

NED, IML, MEH, APP and AV acknowledge the support of NIH/NIAID AI139761. MC and AV acknowledge support from Google Cloud Healthcare and Life Sciences Solutions via the GCP research credits program. MEH acknowledges support from the NIH/NIGMS U54 GM111274 EM acknowledges partial support by MINECO (FIS2016-78904-C3-3-P). YM acknowledges partial support from the Government of Aragon and FEDER funds, Spain through grant E36-17R (FENOL), and by MINECO and FEDER funds (FIS2017-87519-P). AA and YM acknowledge support from Intesa Sanpaolo Innovation Center. The funders had no role in study design, data collection, and analysis, decision to publish, or preparation of the manuscript.

## Authors’ contributions

AA, DMC, MA, AV, EM and YM designed research; AA performed research with contributions from DMC; AA, DMC, MA, AV, EM, and YM analyzed the results. AV and YM wrote the first draft of the manuscript, and all other authors discussed results and edited the manuscript. All authors approved the final version.

## Correspondence

Correspondence and requests for materials should be addressed to AV, EM and YM.

